# Prevalence, knowledge, causes, and practices of self-medication during the COVID-19 pandemic in Bangladesh: A cross-sectional survey

**DOI:** 10.1101/2023.06.27.23291974

**Authors:** Sadia Mahmud Trisha, Sanjana Binte Ahmed, Md Fahim Uddin, Tahsin Tasneem Tabassum, Nur-A-Safrina Rahman, Mridul Gupta, Maisha Samiha, Shahra Tanjim Moulee, Dewan Ibna Al Sakir, Vivek Podder

## Abstract

**Background:** During the COVID-19 pandemic, self-medication (SM) has become a critical element in the healthcare system. SM can ease the burden on hospitals and medical resources by treating minor illnesses. However, inappropriate SM practices can lead to adverse drug reactions, drug resistance, and incorrect diagnoses, resulting in poor health outcomes.

**Methods:** To evaluate the prevalence, knowledge, causes, and practices of SM among the Bangladeshi population during the COVID-19 outbreak, a cross-sectional survey with structured questionnaires was conducted in Chittagong from March to May 2022. The survey included 265 participants, with an average age of 35.09 years, and a multiple-choice questionnaire was used to gather information.

**Results:** The study found that 64.15% of respondents had sufficient knowledge of SM, while 35.8% had insufficient knowledge. The primary reasons for SM during the pandemic were the influence of friends/family (90.74%), fear of infection or contact with COVID-19 cases (73.15%), and fear of quarantine or self-isolation (72.22%). Analgesics/pain relievers (84%) were the most commonly used drugs for SM for COVID-19 prevention and treatment. Antiulcerants/anti acid (42%), Vitamin C and Multivitamin (42%), and Antibiotics (32%) were also frequently used.

**Conclusion:** This study suggests that SM is prevalent among Chittagong City residents, particularly those with less than a tertiary education. The study highlights the importance of building awareness about SM practices and taking necessary steps to control them.

## Background

In January 2020, the World Health Organization (WHO) announced a public health emergency in response to the emergence of COVID-19 [1]. By six months later, approximately 20 million cases and 700,000 deaths had been reported globally [1]. In response to fear of contracting COVID-19, limited access to healthcare services, and misinformation, individuals turned to self-medication (SM). As people were confined to their homes and had limited access to reliable information, the internet became their primary source of information [2]. Moreover, due to crowded hospitals, many individuals opted for SM instead of seeking medical attention.

SM is the behavior of self-treating physical or psychological symptoms based on one’s own perception without consulting a certified physician [2]. Re-use of the previous drug prescribed by the physician, inappropriate usage of over-the-counter drugs (OTC), or purchasing drugs without a prescription is also part of SM [2, 3]. SM is a prevalent practice globally, with a prevalence rate of 32.5% to 81.5% [2, 3]. In Bangladesh, the high cost of treatment, delayed access to healthcare facilities, dissatisfaction with health services, and a low level of specialist proportion are some of the reasons behind the practice. SM has a vital role in our healthcare system. It reduces hospitals’ load and saves medical resources as minor illnesses can be treated with self-prescribed medicines. It also cuts off the cost of treatment and saves time for waiting to see the doctor [3, 4, 5]. However, the act of self-medicating has several negative consequences such as wastage of resources, the development of pathogen resistance, and antibiotic resistance [2, 6]. Furthermore, it is linked to inaccurate dosages, improper administration routes, prolonged use, inadequate drug storage, drug interactions, excessive medication use, and the risk of addiction and abuse [7]. This practice is considered a significant global public health issue due to its potential harmful effects. Most people engage in SM due to a sense of mildness of their symptoms and a belief that they do not require professional medical attention, previous successful self-treatment experiences, the notion of being able to take care of themselves, and limited access to medical care [6, 7].

Since self-treatment involves self-diagnosis, which often leads to errors in diagnosis and treatment choice. Patients may unknowingly take the same active ingredient under different names, leading to double medication or harmful interactions. Incorrect prescription, such as administering a medication intravenously instead of intramuscularly, is also a risk. These factors emphasize the potential dangers of SM and the importance of exercising caution. According to a study in Iran, SM is responsible for 67% of the disease burden worldwide, and studies also show that women who take SM during pregnancy result in 3% of congenital abnormalities [6]. Globally, the prevalence of SM practice is 32.5%-81.5% [4]. It is also a common scenario for Bangladesh. People mainly take SM because of the high cost of treatment, a similar experience with the previous disease, delayed access to the health facility, and dissatisfaction with health services [6, 7, 8]. Another important reason is the low level of persistent specialist proportion. According to a report published by the health ministry, there were six specialists, medical caretakers, and birthing assistants for every 10,000 individuals [7].

During the period of the COVID-19 contagion, the healthcare system faced a massive challenge, especially in developing countries like ours. Periodic lockdown, social distancing, and fear of being contacted by healthcare workers have kept many people from visiting the health facility [7, 8]. With a rapidly increasing number of infected people, Bangladesh’s health system is going through a massive strain with a limited resource setting. Inadequate healthcare workers, lack of sufficient hospital beds, and restrictions in doctors’ visiting hours have forced many people to be ignored by the treatment support they need. Moreover, the nationwide lockdown has affected the economic sector greatly. Many people have lost their livelihood [9].

Azithromycin and doxycycline were the most broadly utilized antimicrobial agents amid the outbreak of COVID-19. Research conducted in Dhaka city shows that ivermectin was also used by 77.15% of the respondents, possibly due to media broadcasts [7]. Vitamin C and multivitamins were also on the top list to be purchased as prevention from COVID-19 [7, 11]. Google search for chloroquine and hydroxychloroquine has increased, indicating that public interest has grown for these drugs presuming them to be the cure for COVID-19 [12]. In Nigeria, the prevalence of SM practice was 41% for the anticipation and treatment of COVID-19 [10]. Similarly, Poland shows a 45.6% prevalence of SM practice, whereas, in Bangladesh, the prevalence was 88.33%, which is very high [7, 8, 9]. All these SM practices can lead to severe health hazards, including drug-induced antimicrobial resistance. The results from this study can evaluate the SM practice among the Bangladeshi population and reveal the factors associated with it so that the policymakers and responsible management can build awareness about SM practice and take the necessary steps to keep it in control.

Therefore, this study aims to evaluate the prevalence and factors associated with SM practice among the Bangladeshi population during the COVID-19 pandemic. The results of this study can provide valuable insights into the extent of SM practice in Bangladesh and identify the factors that contribute to it. Policymakers and health authorities can use these findings to create awareness campaigns and educational programs to encourage safe and appropriate medication use. This study can also help in developing policies to regulate the sale of prescription drugs and over-the-counter drugs to minimize the potential risks associated with SM.

## Methods

Study Design: This study employed a cross-sectional study design to investigate SM practices during the COVID-19 pandemic in Chittagong city, Bangladesh. The study was conducted from March 2022 to June 2022, chosen based on the COVID-19 situation in the country during this period. This period was selected to capture the impact of the pandemic on SM practices in the study population. Convenient sampling was used to select study participants. The researcher visited various areas of Chittagong city and approached individuals who met the inclusion criteria and were willing to participate in the study.

### Target Population

#### Inclusion Criteria

- Participants who have been ill in the past six months
- Residents of Chittagong city
- Residents aged 18 years or above living with their parents
- Non-medical professionals

#### Exclusion Criteria

- Individuals who are unwilling to participate in the study
- People from a medical background
- Respondents with a history of comorbidity
- Respondents practicing SM for more than two years

### Sample Size

The sample size was calculated using the formula: n = (z^2* p*q) / d^2. With a prevalence of SM practices of 60.2% in Savar city, Bangladesh, a margin of error of 0.05, and a confidence level of 95%, the minimum required sample size was calculated to be 369.

### Data Collection Tools

The data collection tool was a structured questionnaire consisting of two sections. The first section collected demographic information, while the second section collected information on SM practices, causes, and reasons using structured KCP scales.

### Data Management and Analysis Plan

After data collection, the accuracy and completeness of all questionnaires were verified to ensure there was no missing or incorrect information. The data were then entered into IBM’s SPSS 25 statistical software for analysis. Descriptive statistics such as frequency distributions, means, and standard deviations were used to summarize the data.

### Quality Control & Quality Assurance

Several quality control and assurance measures were implemented to ensure the reliability and validity of the study results. These measures included regular assistance and direction from the supervisor, reliability checks on the data, and the use of a pre-tested questionnaire for data collection.

### Ethical Consideration

Ethical considerations were taken into account during the study. Participants were informed about the purpose of the study and their right to decline or leave the study at any time.

### Statistical Analysis

Descriptive statistics were used to summarize the characteristics of the study population, including frequency, percentage, mean, and standard deviation. The chi-square test was used to examine the association between demographic characteristics and SM practices. The level of significance was set at p < 0.05.

## Results

### Characteristics of Study Participants and Prevalence of Self-Medication

A total of 265 respondents participated in the survey, with a mean age of 35.09 years (SD=12.45 years). The majority of respondents were female (50.2%). Chi-square test was conducted to examine the association between demographic variables and SM practices. Results showed that males had a prevalence of 43.9% compared to females at 67.5% (p=0.006). Respondents aged 40 years and above had a statistically significant higher prevalence of SM practices at 40.2% (p=0.006). Employment status did not show a statistically significant association with SM practices (p=0.160), nor did level of education (p=0.757). Among respondents, 64.15% had sufficient knowledge about SM practices, while 35.8% had insufficient knowledge. However, those with insufficient knowledge had a significantly higher prevalence of SM practices at 81.7% (p=0.01). [Table 1]

**Table 1:** Characteristics of Study Participants and Prevalence of Self-Medication Practices.

| **Variables** | **Frequency** | **Percentage** | **Prevalence of self-medication** | ***p*-value** |
| --- | --- | --- | --- | --- |
| **Gender** |  |  |  |  |
| Female | 133 | 50.2 | 37.6 | 0.293 |
| Male | 132 | 49.8 | 43.9 |  |
| **Age group** |  |  |  |  |
| <40 | 179 | 67.5 | 40.2 | 0.006 |
| 40-59 | 67 | 25.3 | 50.7 |  |
| >60 | 19 | 7.2 | 10.5 |  |
| **Marital status** |  |  |  |  |
| Married | 166 | 62.6 | 41.6 | 0.939 |
| Single | 84 | 31.7 | 39.3 |  |
| Widowed/divorced | 15 | 5.7 | 40.0 |  |
| **Education** |  |  |  |  |
| 0-8 years | 23 | 8.7 | 47.8 |  |
| 9-12 years | 46 | 17.4 | 41.3 | 0.757 |
| >13 years | 196 | 74.0 | 39.8 |  |
| **Employment** |  |  |  |  |
| Day laborer | 20 | 7.5 | 40.0 | 0.160 |
| Employed | 134 | 50.6 | 47.0 |  |
| Housewife | 40 | 15.1 | 37.5 |  |
| Not-working | 71 | 26.8 | 31.0 |  |
| **Chronic disease** |  |  |  |  |
| No | 203 | 76.6 | 37.9 | 0.09 |
| Yes | 62 | 23.4 | 50.0 |  |
| **Any physician in house** |  |  |  |  |
| No | 217 | 81.9 | 419 | 0.405 |
| Yes | 48 | 18.1 | 35.4 |  |
| **Knowledge** |  |  |  |  |
| Sufficient | 170 | 64.15 | 12.2 |  |
| Insufficient | 95 | 35.85 | 81.7 |  |

### Causes for Self-Medication of COVID-19

A multiple-choice questionnaire was administered to determine the reasons for SM among respondents. The results were presented in a bar graph. The top reasons for SM for COVID-19 were influence from friends/family (90.74%), fear of infection or contact with a suspected or known COVID-19 case (73.15%), fear of quarantine or self-isolation (72.22%), unavailability of drugs for COVID-19 treatment in health facilities (62.04%), and delay in receiving treatment at health facilities (41.67%). Other reported reasons included influence from social media (25.93%). [Figure 1]

**Figure 1:**
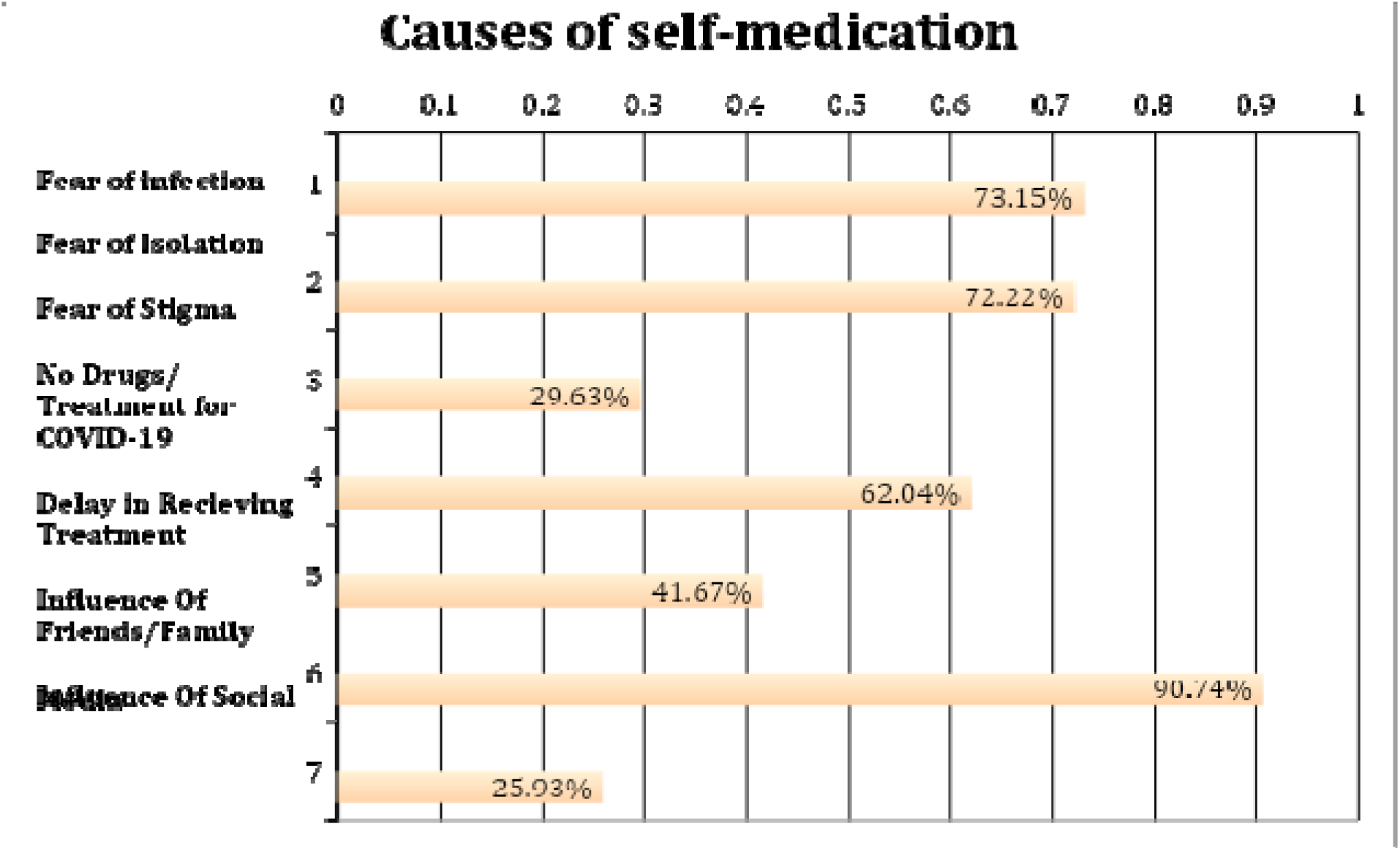
Causes for Self-Medication for COVID-19 among Study Participants (n=265)

### Causes for Self-Medication in the Treatment and/or Prevention of COVID-19

The pie chart illustrates the factors behind SM. The majority of participants (66%) reported emergency illness as the primary cause of SM. Proximity to the pharmacy was the second most cited factor (20%). Other reasons included delayed access to hospital services (9%) and cost of health facilities (5%). [Figure 2]

**Figure 2:**
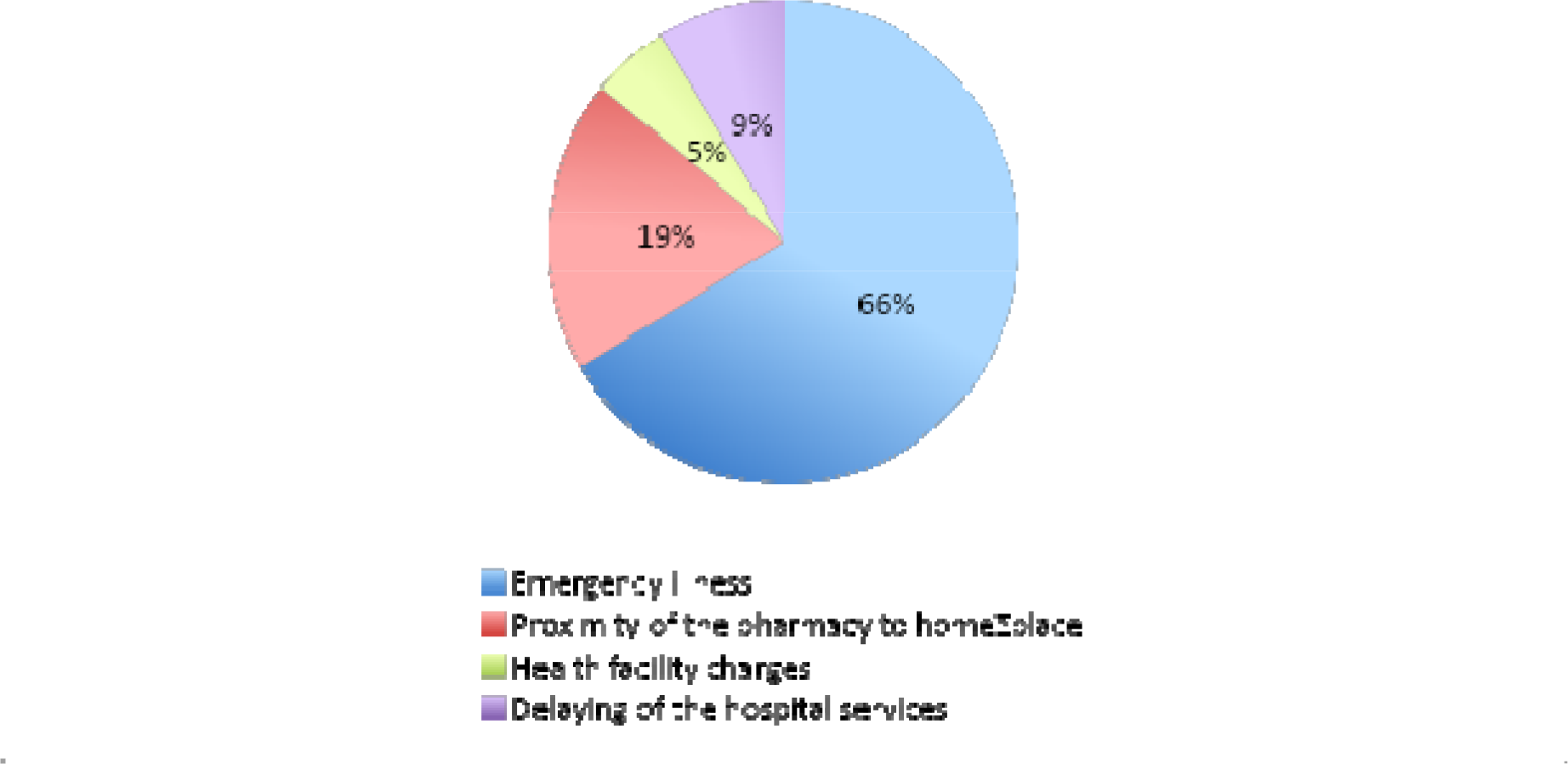
Causes of Self-Medication Practices for the Treatment and/or Prevention of COVID-19 as reported by respondents.

Based on Figure 3, the most commonly used drugs for SM in the treatment and prevention of COVID-19 were antipyretics/analgesics at 87%, followed by Vitamin C and multivitamins at 45%, antacids at 32%, antibiotics at 30%, and herbal products at 25%. Antitussives were used by 24% of respondents, while sedatives were the least used drugs at 5%.

**Figure 3:**
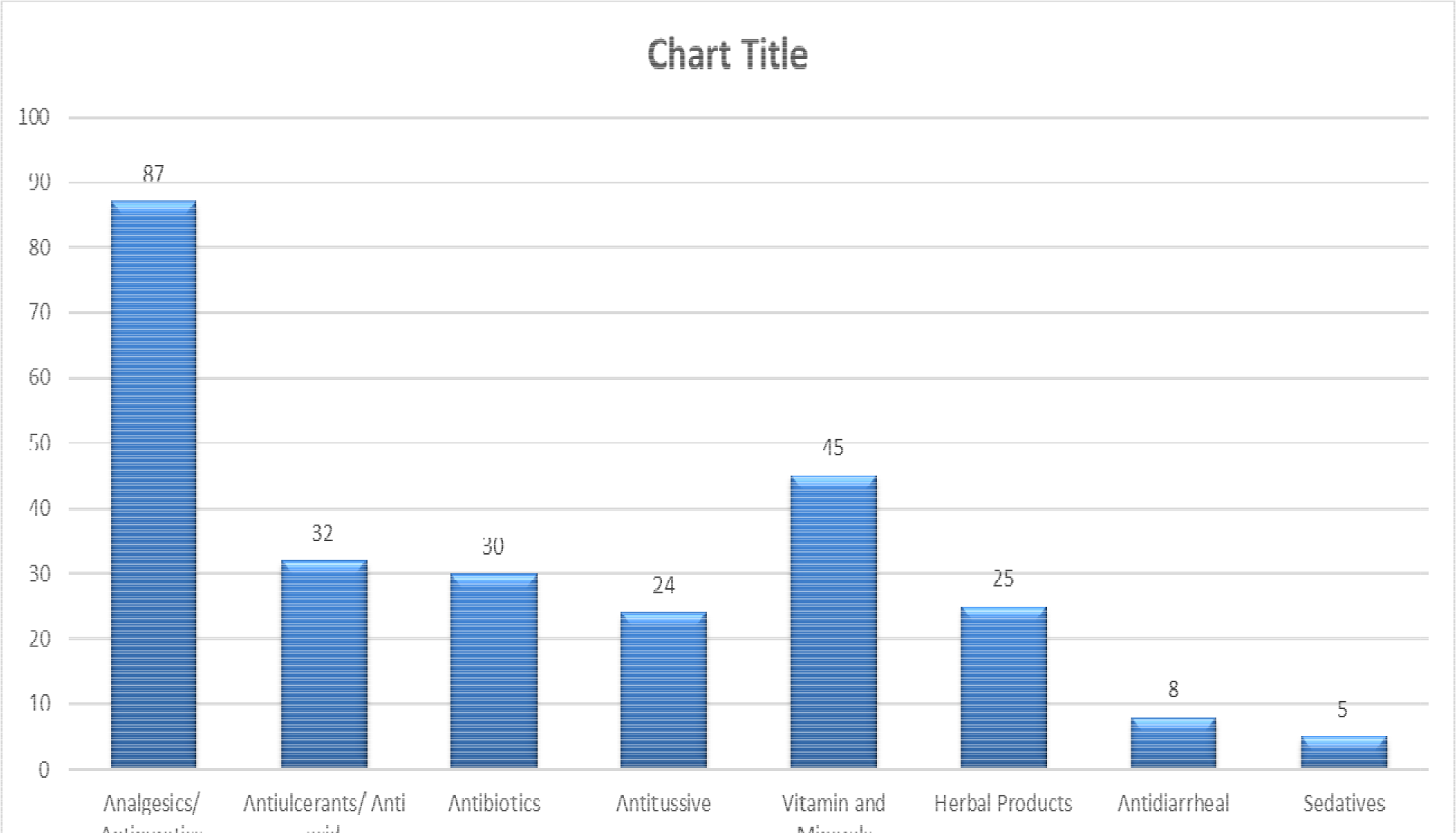
Types of drugs used for self-medication in the treatment and prevention of COVID-19 among respondents (n=265)

### Multivariate logistic regression analysis of factors associated with self-medication practices for COVID-19

The multivariate logistic regression model revealed that males had 1.05 times higher odds of practicing SM compared to females [OR=1.05, 95% CI: 0.57-1.96]. The odds of practicing SM among respondents aged >60 years were 92% lower compared to those below <40 years [OR: 0.08, 95% CI: 0.01-0.68]. Furthermore, the table 3 shows that respondents in the 40-59 years age group, single, employed, housewives, not working, and those with chronic diseases had higher odds of practicing SM compared to married, day laborers, and those without chronic diseases.

**Table 3:** Association of self-medication practice with sociodemographic characteristics of participants:

| **Variables** | **AOR (95% CI)** |
| --- | --- |
| **Gender** |  |
| Female | Reference |
| Male | 1.05 (0.57 - 1.96) |
| **Age group** |  |
| <40 | Reference |
| 40-59 | 1.41 (0.70 - 2.83) |
| >60 | 0.08 (0.01 - 0.40) |
| **Marital status** |  |
| Married | Reference |
| Single | 1.12 (0.51 – 2.47) |
| Widowed/divorced | 0.91 (0.26 – 3.01) |
| **Education** |  |
| 0-8 years | Reference |
| 9-12 years | 0.31 (0.04 – 1.64) |
| >13 years | 0.20 (0.03 – 1.02) |
| **Employment** |  |
| Day laborer | Reference |
| Employed | 8.67 (1.42 – 34.35) |
| Housewife | 6.95 (1.09 – 21.60) |
| Not working | 4.72 (0.74 – 22.51) |
| **Chronic disease** |  |
| No | Reference |
| Yes | 2.71 (1.35 – 5.61) |
| **Any physician in house** |  |
| No | Reference |
| Yes | 1.74 (0.35 – 1.50) |

## Discussion

We examined the knowledge, prevalence, predictors, and causes of SM for COVID-19 prevention and treatment among individuals in Chittagong City, Bangladesh. Unlike previous studies that relied on online surveys of literate populations, our study included participants from diverse socio demographic backgrounds.

Our study found that SM practices were equally prevalent among male and female participants, with the majority belonging to the age group of <40. This may be more likely because young adults engage in risky behavior and are less likely to comply with health recommendations However, we also observed a higher prevalence of SM among the 40-59 age group, which is similar to a study conducted in Dhaka City [7]. Furthermore, our study found that people’s knowledge, causes, and determinant factors had a significant effect on SM practices to fight COVID-19 [8]. This suggests that targeted education campaigns may be an effective strategy for improving compliance with SM measures. Interestingly, we also found that the prevalence of SM was higher among people with insufficient knowledge, consistent with a study conducted in Nigeria [11]. This highlights the importance of providing accurate and accessible information about the risks and benefits of self-medication, as well as promoting safe and effective healthcare practices.

Our study found that a significant proportion of participants engaged in self-medication for COVID-19 prevention and treatment, with 40.8% reporting this practice. While this percentage is lower than previous studies conducted in Dhaka City [10], it is still a cause for concern given the potential risks and negative health outcomes associated with self-medication. Furthermore, we identified that middle-aged individuals, those who were married, had an education level below tertiary, and those with insufficient knowledge about SM were more likely to engage in SM practices, similar to findings from studies conducted in Nigeria and Dhaka City [7, 11]. This may be due to a lack of access to healthcare resources or a perception that self-medication is a more convenient or cost-effective option. Self-medication during the pandemic can worsen existing health crises for which countries are unprepared. Restricted media announcements, involvement of pharmacists and drug regulators, and support from national health authorities can help mitigate the risks of self-medication, drug shortages, and price hikes [12, 13, 14].

Our study also revealed that the reasons for SM included fear of infection, isolation, and stigma, influence of friends and family, delay in receiving treatment, unavailability of COVID-19 drugs, and social media. Emergency illness, proximity to a pharmacy, and health facility charges were also cited as reasons for SM. COVID-19 fears and stigma, as well as social media and peer pressure, increase SM use as people avoid seeking medical treatment in hospitals or clinics and may follow inaccurate information online [15, 16, 17]. It is worth noting that our findings contrast with those of previous studies conducted in Savar and Dhaka City. The differences in the reasons for SM may reflect the evolving circumstances of the pandemic and the varying attitudes and behaviors of individuals towards seeking medical care [6, 7].

Most participants used analgesics/antipyretics for COVID-19 prevention and treatment, followed by vitamins/minerals, likely due to their immunity-boosting effects and over-the-counter availability. Also, antiulcerants, antitussives, herbal products, and antibiotics were used, with 32% using antibiotics, possibly due to social media and peer influence. These findings agree with prior studies that reported the widespread use of azithromycin and vitamins/minerals for COVID-19 [6, 7, 11].

Pharmacies were the primary source for purchasing medicines, possibly due to proximity and a lack of regulation that allows pharmacy workers to sell medicines without prescriptions. This finding is consistent with a study conducted in Nigeria [17]. Our study sheds light on the prevalence, predictors, and causes of SM practices for COVID-19 prevention and treatment in Dhaka City, Bangladesh. Our findings highlight the importance of increasing knowledge about appropriate SM practices and the need for regulations to prevent the sale of prescription drugs without a prescription [18, 19].

## Limitations

Despite its strengths, this study had some limitations. First, we did not collect information on drug doses or duration of treatment, which limits our ability to draw conclusions about the effectiveness and safety of the medications used. Additionally, we did not verify the quality or authenticity of the herbal products used as traditional medicines, which could have impacted the results. Another limitation is that our study was conducted in Chittagong city and may not be generalizable to other regions in Bangladesh. The small sample size of our in-person surveys also limits the generalizability of our findings. Finally, as the study relied on self-reported data, social desirability bias may have influenced the observed prevalence of SM for alleged COVID-19 prevention and/or treatment. Despite these limitations, this study provides valuable insights into the knowledge, factors, behaviors, and potential predictors of SM among Chittagong residents of Bangladesh, and the face-to-face interviewing approach used for the investigation in a short amount of time is a strength of the study.

## Conclusion

This study provides valuable insights into the prevalence, predictors, and reasons for SM among residents of Dhaka city, Bangladesh during the COVID-19 pandemic. Our findings suggest that a significant proportion of the population engaged in SM for prevention and treatment of COVID-19, with common drugs including analgesics, vitamins, antiulcerants, and antibiotics. Lack of knowledge about SM and fear of infection and isolation were identified as significant predictors of SM. Our study also highlights the need for increased awareness campaigns to counter incorrect information on social media and better regulation to prevent pharmacies from selling medicines without a prescription. Overall, these findings have important implications for public health efforts to address the COVID-19 pandemic in Bangladesh and other similar settings.

## Data Availability

All data produced in the present study are available upon reasonable request to the authors.

## Declarations

### Ethics approval and consent to participate

The study followed the ethical principles set forth in the Declaration of Helsinki and was granted approval by the ethics committee at North South University in Dhaka, Bangladesh. Before the data collection, participants provided written informed consent.

### Consent for publication

Not applicable

### Availability of data and materials

The datasets generated and/or analysed during the current study are not publicly available but are available from the corresponding author on reasonable request.

### Authors’ contributions

Conceptualization: SMT, SBA, MFU; Data curation: SMT, SBA, TTT; Formal analysis:: SMT; Investigation:: SMT; Methodology:: SMT, SBA, MFU, NSR, MG; Project administration:: SMT, SBA, MFU, MG, MS, STM; Software:: SMT; Supervision:: SMT; Validation:: SMT, SBA, MFU; Visualization; Roles/Writing - original draft:: SMT, SBA, MFU, TTT, NSR, MG, STM, DIAS; Writing - review & editing: SMT, SBA, MFU, TTT, NSR, MG, STM, DIAS.

## Acknowledgements

Not applicable

## List of Abbreviations

SM: Self-medication
WHO: World Health Organization
OTC: Over-the-counter drug

